# Insomnia, excessive daytime sleepiness, anxiety, depression and socioeconomic status among customer service employees in Canada

**DOI:** 10.1101/19003194

**Authors:** FA Etindele Sosso

## Abstract

**Objectives:** The present study alerts on the potential effect of working full time in a call center as a risk factor for neuropsychiatric illnesses. It is the first study investigating deeply presence of anxiety, depression, insomnia and excessive daytime sleepiness among a large population of customer service employees. It has 3 specifics goals which were (1) document presence of sleep disorders among customer service advisors (2) document presence of anxiety and depression in this population (3) determine the influence of the socioeconomic status, duration in position and full time or part-time shift on the diseases above.

**Findings:** It was found that the majority of people working in customer service are undergraduate students or at a secondary/high school degree. They worked full time, are single and have reported at least two of the neuropsychiatric disorders assessed in the present study. Among customer service advisors enrolled in this study, all neuropsychiatric disorders investigated were present and significantly higher for those working full time. Perceived socioeconomic status (pSES) was almost similar for full time and part time workers with a mean score of 4.8 on the MacArthur scale of subjective social status. Results revealed that duration in position was an excellent predictor of insomnia, sleepiness and anxiety (respectively with R2=91,83%, R2=81,23% and R2=87,46%) but a moderate predictor of depression (R2=69,14%). The pSES was a moderate predictor of sleep disorders (respectively R2=62,04% for insomnia and R2=53,62% for sleepiness) but had a strong association with anxiety and depression (R2=82,95% for anxiety and R2=89,77% for depression). It was found that insomnia and anxiety are more prevalent for immigrants and international students compared to Canadians, while depression was similarly higher for Canadian and immigrants compared to international students. It was found that sleepiness has the same trend in the three subgroups.

**Conclusion:** Customer service employees are exposed to a continuous stimulation of their cognitive functions in addition to different stressors which can progressively and silently damage the nervous system. Investigations on mental and physical health of customer service advisors are worthy of interest, and understanding how their work, their rotating shifts and their socioeconomic status influence their resilience and their performance at work; may help comprehension of similar health issues emerging in similar populations with similar occupations.

## 1. INTRODUCTION

Nowadays, accumulating literature suggests that our modern lifestyle is accompanied by many stressors such as night work, continued flow of news via social network, a low socioeconomic status complicating healthcare or, more often, successive bad experiences leading to anxiety or depression (Arman et al., 2011; Assari et al., 2016). An important part of this modern age is the possibility to complete almost everything online or by phone. Even if this evolution makes our lives easier, evidence suggests that cell phones have a negative impact on our physical health, our socialization and our brains (Wei et al., 2012; Arora et al., 2013a; Etindele Sosso and Raouafi, 2016; 2017; Etindele Sosso, 2018). Many studies reported associations between lack of physical activity and obesity (Arora et al., 2013a; Arora et al., 2013b), between poor and moderate physical activity and cardiovascular diseases (Nicholson et al., 2012; Etindele Sosso and Raouafi, 2016), or development of cognitive impairment with an excessive use of technology (Shams et al., 2015). The impact of this lazy lifestyle on the consumer’s body has now been well documented, but there is little to no data available regarding the health of a few main actors of this new age: the advisors. Whatever fields you are working in, be it shopping, technical support, shopping, need technical support, ordering, or just seeking some help or information; there are human beings behind the screen or the phone. Recent research in chronobiology and sleep medicine reported incidence of some type of work on mental health (Etindele Sosso, 2017a; b). For example, night shift workers and pilots are exposed to jetlag (Jagannath et al., 2013; Kalmbach et al., 2015). Nurses and rotating shift workers who have constant flux in their shifts face a permanent disturbance of their circadian system, experience premorbid psychobiological processes as well as work-related depression and anxiety (Kalmbach et al., 2015). Owing to the permanent flux of rotating shifts in these types of jobs, work-related sleep-wake schedules often conflict with internal circadian rhythms (Kalmbach et al., 2015). Sleep-wake impairments in response to a circadian challenge are highly variable from an individual to another, but an important proportion of shift workers cannot synchronize their circadian clocks to their work-related sleep-wake schedules. Indeed, up to 26% of rotating shift workers develop shift work disorder, which is characterized by persistent and severe sleep disturbance during the sleep period and/or excessive sleepiness during the wake period (Jagannath et al., 2013; Kalmbach et al., 2015). The area of customer service is remarkably similar to those of nursing and night shift workers in terms of rotating shifts, night shift work and volume of services the advisors have to provide. Regardless of the domain where an advisor is providing the customer service, responding to so many calls without break during several hours and often with a significant speed probably takes a toll on the advisor’s mental health after a while. The problematic is the fact that this topic was not deeply investigated, perhaps non explored at all. This was confirmed by the difficulty to find literature on this subject in the most popular databases such as PubMed, Scopus and Web of Science. Currently, there is no data on the prevalence of sleep disorders or what kind of sleep disorders customer service advisors’ experience during their shifts or because of this job. Similarly, there is no literature on mood disorders related to working in customer service. The present paper is an exploratory study which has as objectives to (1) document presence of sleep disorders among customer service advisors (2) documented presence of anxiety and depression in this population (3) and determine the influence of the socioeconomic status, duration in position and full time or part-time shift on the diseases above.

## 2. MATERIALS AND METHODS

### 2.1 Ethical Statement

This project was part of a larger study approved initially by the ethics committee of research on humans of the Université de Montreal in Canada, and secondly by the team leaders or human resources officer of all the call centers and customer services which provide advisors. Data were collected and computed in an external database. This database was secured outside the public network and was shared only with the investigators involved in the project.

### 2.2 Participants and procedure

The customer service’s call centers were contacted during the national career event and the national job fair, which both took place in three Canadians cities (Montreal, Laval and Longueil) in 2016 and 2017. Study objectives and protocol were explained to the representatives on-site and an electronic link to complete the study online was provided during the event. Two reminders to participate in the study were sent by email (two days and one week after the event) to the representatives we met. This allows us to verify that their companies’ managers approved the study and shared our electronic link with their customer service advisors. A total of 52 customers services were contacted during the events above, but only 11 customer services confirmed their participation to the study (response rate of 21.15%). All participants (n=1238, 640 females and 598 males) received the study link from their administration (from theirs team leaders or supervisors at work). Two contact numbers and email address to reach our research team and the ombudsman were provided in the first page of the study online, following a description of the study in French and English on the same page. The online questionnaire included a consent statement related to data protections and understanding of the study objectives, which required an acceptance and willingness to continue with the anonymous, online survey by clicking “I agree”; if participants wished to move forward with the study. The average time to answer all questions was estimated during our final testing to be thirty minutes. The questionnaire was configured in a way such it could only be completed once by participants with the same email address. Responses were collected between November 2016 and October 2017. Duplicates and incomplete forms were removed (n=38). The final sample analyzed was n=1200 participants/advisors (610 females and 590 males)

### 2.3 Measures

#### Insomnia Severity Index (ISI)

This questionnaire is widely used to assess insomnia and associated parameters, including lack of sleep and sleep quality. It was developed by Morin and al. in 2001 and widely validated and used for different populations and contexts (Bastien et al., 2001; Thorndike et al., 2011). The internal consistency of the ISI was excellent (Cronbach’s α = 0.92), and each individual item showed adequate discriminative capacity (r = 0.65-0.84). The area under the receiver operator characteristic curve was 0.87 and suggested that a cut-off score of 14 was optimal (82.4% sensitivity, 82.1% specificity, and 82.2% agreement) for detecting clinical insomnia. An ISI score between 0 and 7 is an insomnia considered clinically non-significant, between 8 and 14 as a subthreshold insomnia, between 15 and 21 as a moderate clinical insomnia, and between 22 and 28 like as a severe clinical insomnia (Bastien et al., 2001; Thorndike et al., 2011).

#### Epworth Sleepiness Scale (ESS)

The Epworth Sleepiness Scale is a simple and reliable instrument that has been used worldwide since 1991. This test was developed by Dr. Murray Johns of Epworth Hospital in Melbourne, Australia (Johns, 1991). This test assesses drowsiness levels throughout the day, also known as excessive daytime sleepiness. This test also indicates if a certain degree of drowsiness may warrant a visit to the doctor and a more thorough assessment to diagnose a sleep respiratory disorder. The test consists of eight questions (0 to 3 points) and is completed in less than five minutes. If the participant scores 9 points or fewer, it is considered normal. If the participant scores 10 points or more, an appointment should be made with a sleep specialist. The Epworth scale only quantifies subjective drowsiness (interpreted by the participant) regardless of its association with a sleep disorder.

#### The Hospital Anxiety and Depression Scale (HADS)

This is a self-administered scale including 14 items, divided into two subscales of seven items (Anxiety or HADS-A, Depression or HADS-D). It contains no somatic items that can be confused with the symptomatic manifestations of a disease. Each item is scored on a scale of 0 to 3. A score is generated for each of the two sub-scales and for the entire HADS (HADS-T). The scoring is similar for anxiety and depression and following scores, there are three categories of symptomatic levels: non-cases or asymptomatic ones (scores ≤ 7); probable or borderline cases (scores 8-10); clearly or clinically symptomatic cases (scores ≥ 11). The duration of administration is approximately five minutes and psychometric properties are detailed in previous studies (Bjelland et al., 2002; Roberge et al., 2013).

#### The MacArthur Scale of Subjective Social Status

This scale measured in this study the perceived Socioeconomic Status (pSES) with *the MacArthur Scale of Subjective Social Status*. This scale allows a self-report of pSES using the general socioeconomic markers such as income, occupation and education (Giatti et al., 2012; Shaked et al., 2016). The online form showed participants a ladder with a number on each step, ranging from 1 to 10 (lowest to highest). Each participant was asked to choose a step of the ladder corresponding to their current feeling about their social status at this level in their lives. Participants in the present study endorsed values ranging from 2 to 7. Previous studies demonstrated that pSES is an excellent predictor of health outcomes even after adjustment for objective SES measures (Giatti et al., 2012; Shaked et al., 2016).

In addition to sociodemographic data like age, marital status, immigration status (citizen, international student or immigrants), education, duration in position (ranging from 1 month to 24 months in the sample) and sex, participants were classified in full time workers (from 30h/week to 40h/week) and part-time workers (from 20h/week to 29h/week); according to Canadian laws.

### 2.4. Data analysis

The normal distribution of the data was analyzed using a Shapiro Wilk test. Unpaired sample t-tests were used to compare difference in insomnia, sleepiness, anxiety and depression for male vs. female; and for part-time workers vs. full time workers. First, linear regression analyses were used to investigate whether there was association between *insomnia, sleepiness, anxiety, depression* as dependent variables; and *duration in position* as an independent variable. Next, linear regression analyses were used to assess if *insomnia, sleepiness, anxiety and depression* could be predicted with *perceived socioeconomic status* as an independent variable. Finally, a one-way ANOVA with a Tukey multiple comparisons of means was performed to analyze differences between citizens, international students and immigrants in their level of anxiety, depression, insomnia and excessive daytime sleepiness. All non-significant analysis was not reported. All analysis was controlled for age and sex. All the statistical tests used an alpha of 0.05 as level of significance. Data analysis was performed using PRISM (Graph Pad Prism, version 7.0.0.159, Graph pad software) and SPSS version 22 (IBM Corp, Armonk, NY, USA)

## 3. RESULTS

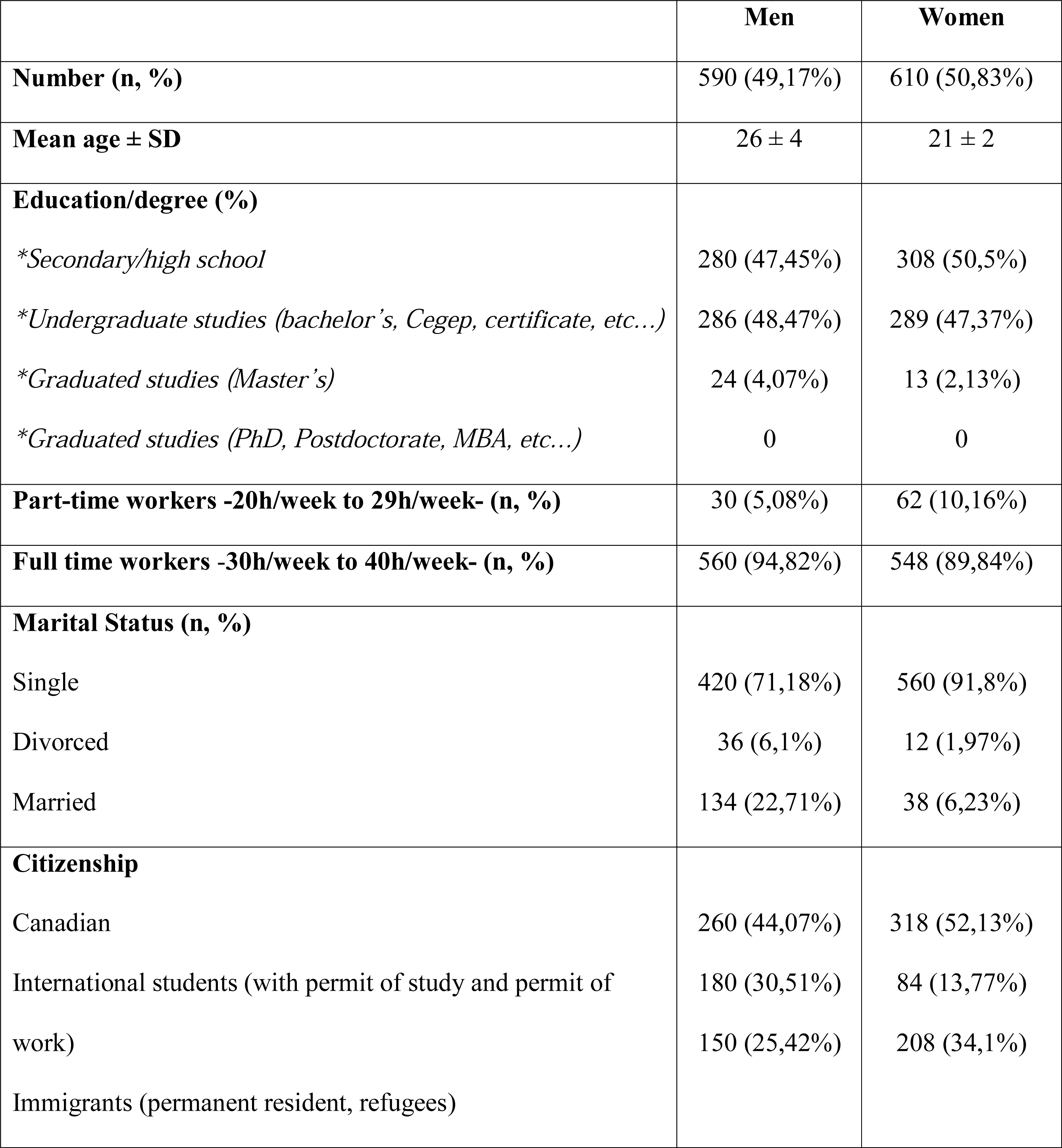

|  | **Men** | **Women** |
| --- | --- | --- |
| **Number (n, %)** | 590 (49,17%) | 610 (50,83%) |
| **Mean age ± SD** | 26 ± 4 | 21 ± 2 |
| **Education/degree (%)** |  |  |
| *\*Secondary/high school* | 280 (47,45%) | 308 (50,5%) |
| *\*Undergraduate studies (bachelor's, Cegep, certificate, etc...)* | 286 (48,47%) | 289 (47,37%) |
| *\*Graduated studies (Master's)* | 24 (4,07%) | 13 (2,13%) |
| *\*Graduated studies (PhD, Postdoctorate, MBA, etc...)* | 0 | 0 |
| **Part-time workers -20h/week to 29h/week- (n, %)** | 30 (5,08%) | 62 (10,16%) |
| **Full time workers -30h/week to 40h/week- (n, %)** | 560 (94,82%) | 548 (89,84%) |
| **Marital Status (n, %)** |  |  |
| Single | 420 (71,18%) | 560 (91,8%) |
| Divorced | 36 (6,1%) | 12 (1,97%) |
| Married | 134 (22,71%) | 38 (6,23%) |
| **Citizenship** |  |  |
| Canadian | 260 (44,07%) | 318 (52,13%) |
| International students (with permit of study and permit of work) | 180 (30,51%) | 84 (13,77%) |
| Immigrants (permanent resident, refugees) | 150 (25,42%) | 208 (34,1%) |

Table 1 presented the descriptive data and the sociodemographic characteristics of the sample. The sample is almost identical in proportion of men and women (respectively, 49.17% and 50.83%), with the majority of them having a secondary/high school degree (47.45% for men, 50.5% for women) and an undergraduate diploma (48.47% for men, 47.37% for women). The Majority of customer service employees in this population were full time workers (94.82% for men and 89.84% for women). The majority of customer service employees reported a marital status ‘‘single’’ (71.18% for men, 91.8% for women), while 22.71% of men were married compared to only 6.23% of women. Another important discovery of this study is the fact that, even customer service employees were mostly Canadian (44.07% of men, 52.13% of women); the other half of workers were immigrants and international students. It was found that in men, 30.51% were international students (both with permits of study or permits of work) and 25.42% were immigrants (those with the status of permanent residents or refugees). Similar trends were found for women with 13.77% of international students and 34.1% of immigrants. Unpaired t tests were used to assess differences in the level of insomnia, excessive daytime sleepiness, anxiety, depression and pSES between full time workers and part-time workers; and between male and female. The figure 1 reported these results. It was found for full time workers an insomnia’s mean score of 17 for men and 16 for women (p < 0.001); and for part-time workers an insomnia’s mean score of 12/28 for men and 15 for women (p < 0.001). Excessive daytime sleepiness score was an average of 9 for both men and women working full time (p > 0.05), and for part-time workers it was found 6 for women and 5 for men (p < 0.001). The figure 1C showed an anxiety’s mean score of 12.5 for men and 11.8 for women working full time (p < 0.001); and 9.8 for men and 9 for women working part-time (p < 0.001). Figure 1D showed similar mean score of 9.8 for both men and women working full time (p > 0.05), and a 9 for both men and women working part-time (p > 0.05). The Figure 1E showed an average pSES of 4.9 for both men and women working full time (p > 0.05), and 4.5 for men and women working part time (p > 0.05). The figure 2 depicts the association between each neuropsychiatric disorders and the duration in position for each employees, assessed with linear regressions. There is strong statistical association between insomnia and duration in position (R^2^=91,83%), excessive daytime sleepiness and duration in position (R^2^=81,23%) and anxiety and duration in position (R^2^=87,46%). The relation between depression and duration in position was moderate (R^2^=69,14%). Figure 3 showed the prediction of the same neuropsychiatric diseases above by the socioeconomic status (pSES). It was found in pictures 3A and 3B, a moderate relation between insomnia and pSES (R^2^=62,04%) and excessive daytime sleepiness and pSES (R^2^=53,62%). The pictures 3C and 3D showed a strong association between anxiety and pSES (R^2^=82,95%) and between depression and pSES (R^2^=89,77%). Figure 4 reported the results of the ANOVA performed to figure out differences between citizens, international students and immigrants in their level of anxiety, depression, insomnia and excessive daytime sleepiness. Figures 4A and 4C showed that being an immigrant is associated with clinical insomnia and more strongly with higher levels of anxiety, followed by the status of international student and lesser by Canadian born (respectively F=40.26, p < 0.001; Post Hoc, p < 0.001 and F=16.27, p < 0.001; Post Hoc, p < 0.001). Figure 4D showed a higher score of depression for Canadians and immigrants compared to international students (F=3.166, p < 0.001; Post Hoc, p < 0.001). Analysis did not find a significant difference in excessive daytime sleepiness for the three sub-groups (F=6; p=0.09 > 0.05; Post Hoc, p < 0.001).

**Figure 1:**
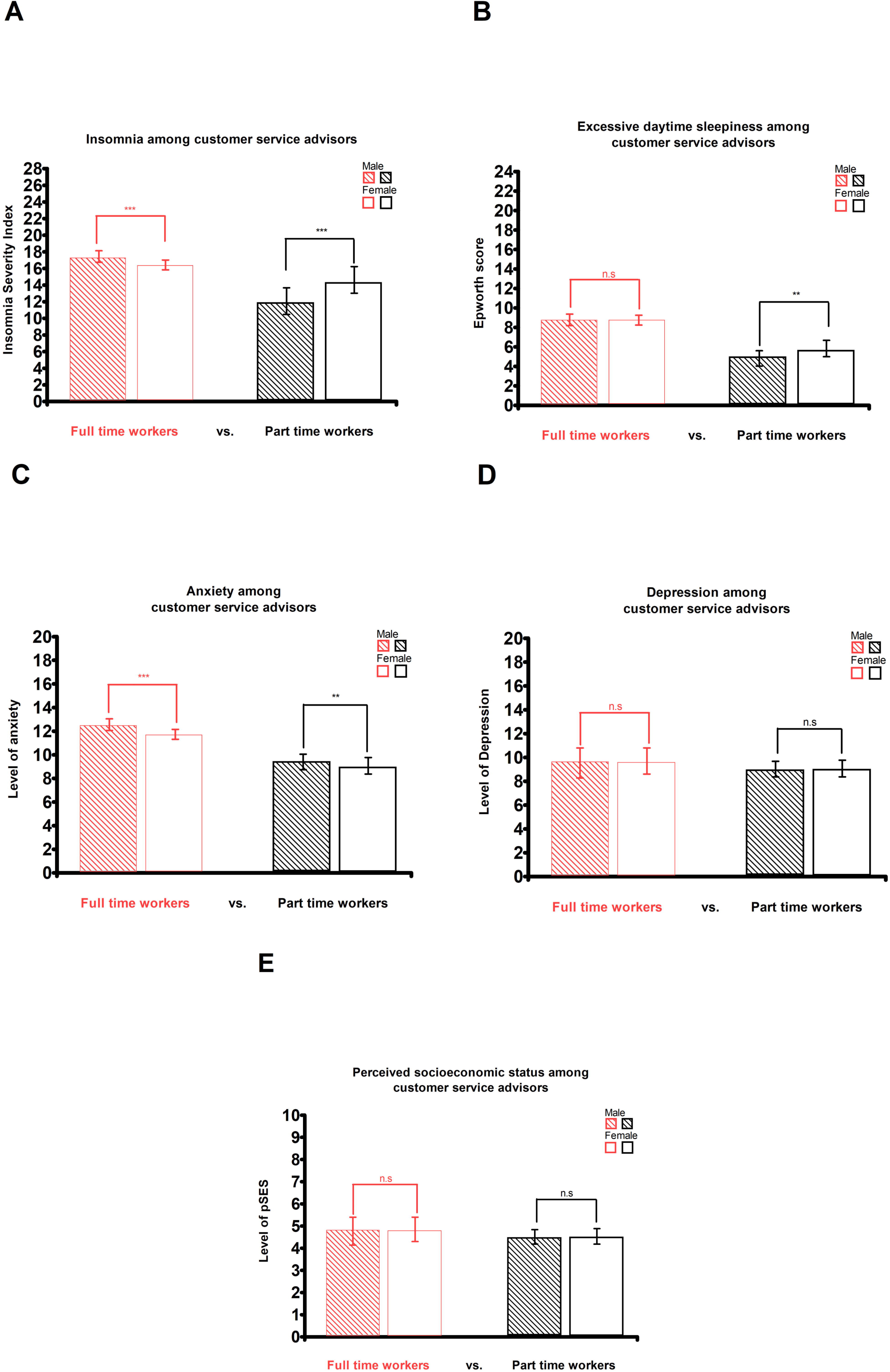
Distribution of neuropsychiatric disorders in the sample. **A: Differences in insomnia’s level among customer service advisors**. Insomnia’s level is the mean of individuals score on the Insomnia Severity Index (ISi) for both men vs. woman and full time workers vs. part time workers. **B: Differences in Excessive daytime sleepiness among customer service advisors**. Drowsiness’s level is the mean of individuals score on the Epworth Sleepiness Scale (ESS) for both men vs. woman and full time workers vs. part time workers. **C: Differences in anxiety among customer service advisors**. Anxiety’s level is the mean of individuals score on the Hamilton anxiety scale (HADS-A) for both men vs. woman and full time workers vs. part time workers. **D: Differences in Depression among customer service advisors**. Depression’s level is the mean of individuals score on the Hamilton depression scale (HADS-B) for both men vs. woman and full time workers vs. part time workers. **E: Differences in socioeconomic status among customer service advisors**. The pSES (perceived socioeconomic status) is based on the mean of individuals score on the MacArthur Scale of Subjective Social Status, for both men vs. woman and full time workers vs. part time workers. Differences were tested with paired t-tests. **Legends:** p < 0.0001 (***), p < 0.001 (**), p < 0.01 (*), p 3 0.05 (n.s)

**Figure 2:**
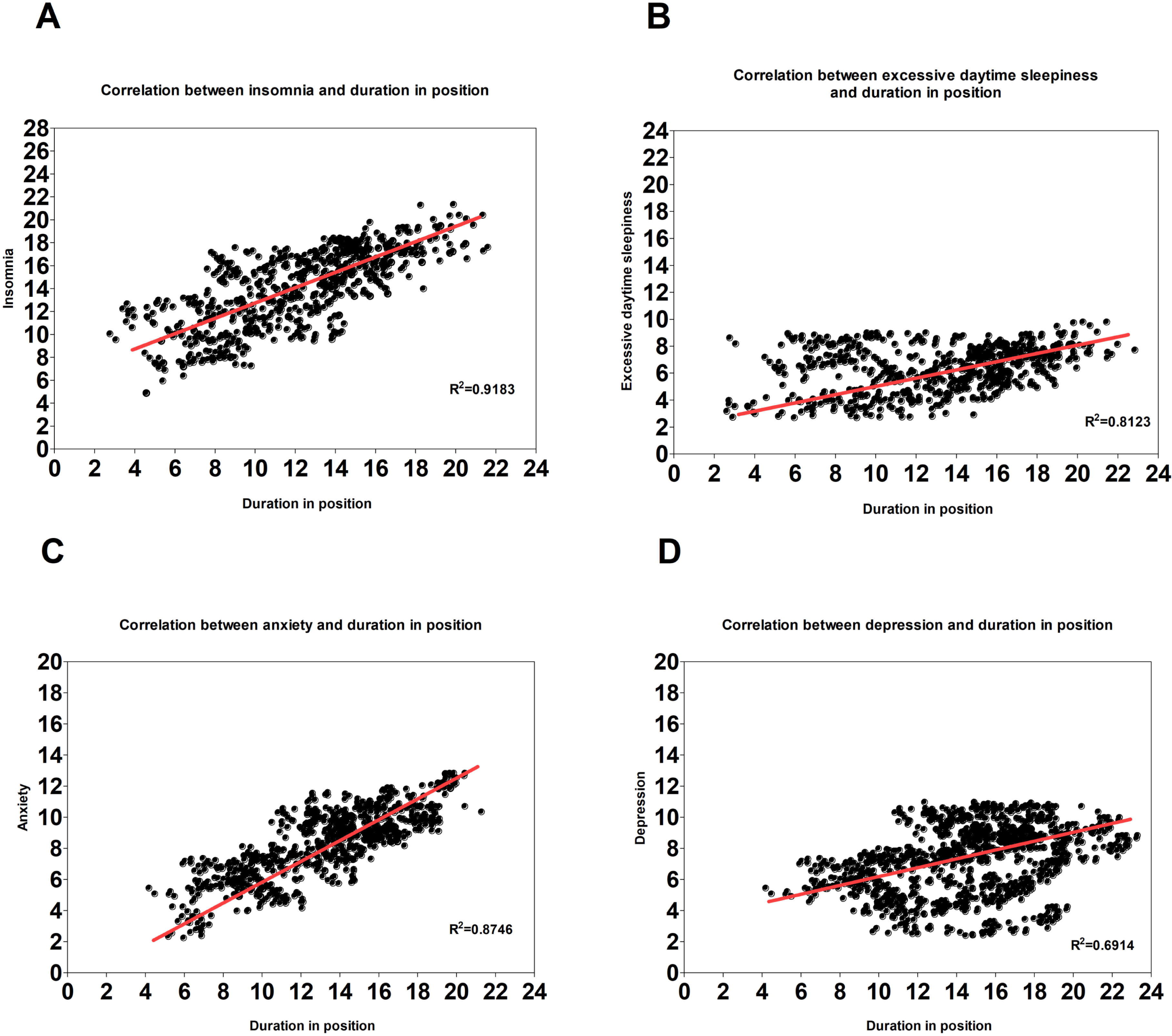
Prediction of neuropsychiatric disorders by the duration in position. Linear regressions were used to assess the correlation between *insomnia, sleepiness, anxiety, depression* as dependent variables; and *duration in position* as an independent variable. All analysis were controlled for age and sex. All the statistical tests used an alpha of 0.05 as level of significance. Confidence interval is 95%.

**Figure 3:**
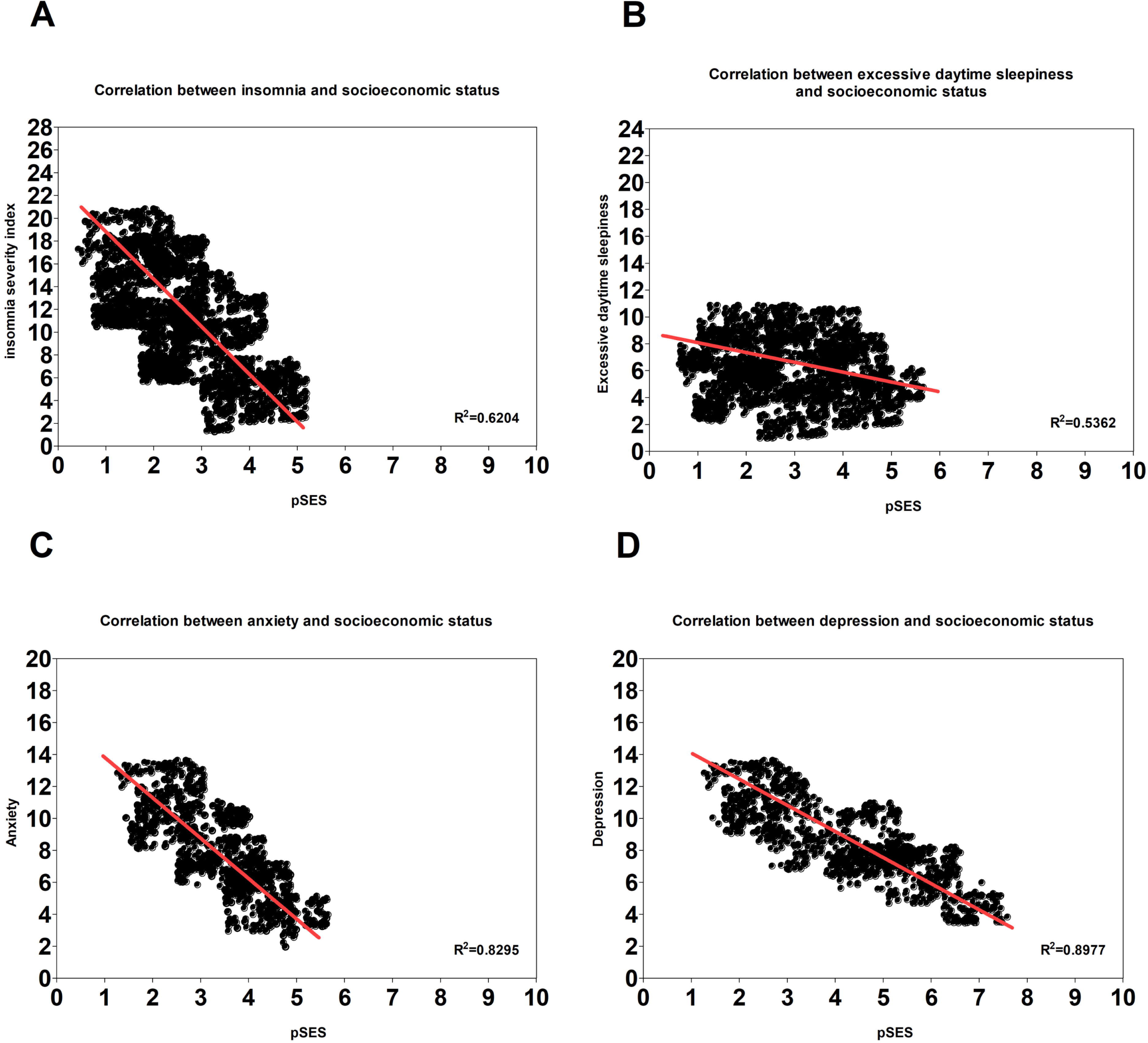
Prediction of neuropsychiatric disorders by the socioeconomic status. Linear regressions were used to assess the correlation between *insomnia, sleepiness, anxiety, depression* as dependent variables; and *perceived socioeconomic status* as independent variable. All analysis were controlled for age and sex. All statistical tests used an alpha of 0.05 as level of significance. Confidence interval is 95%.

**Figure 4:**
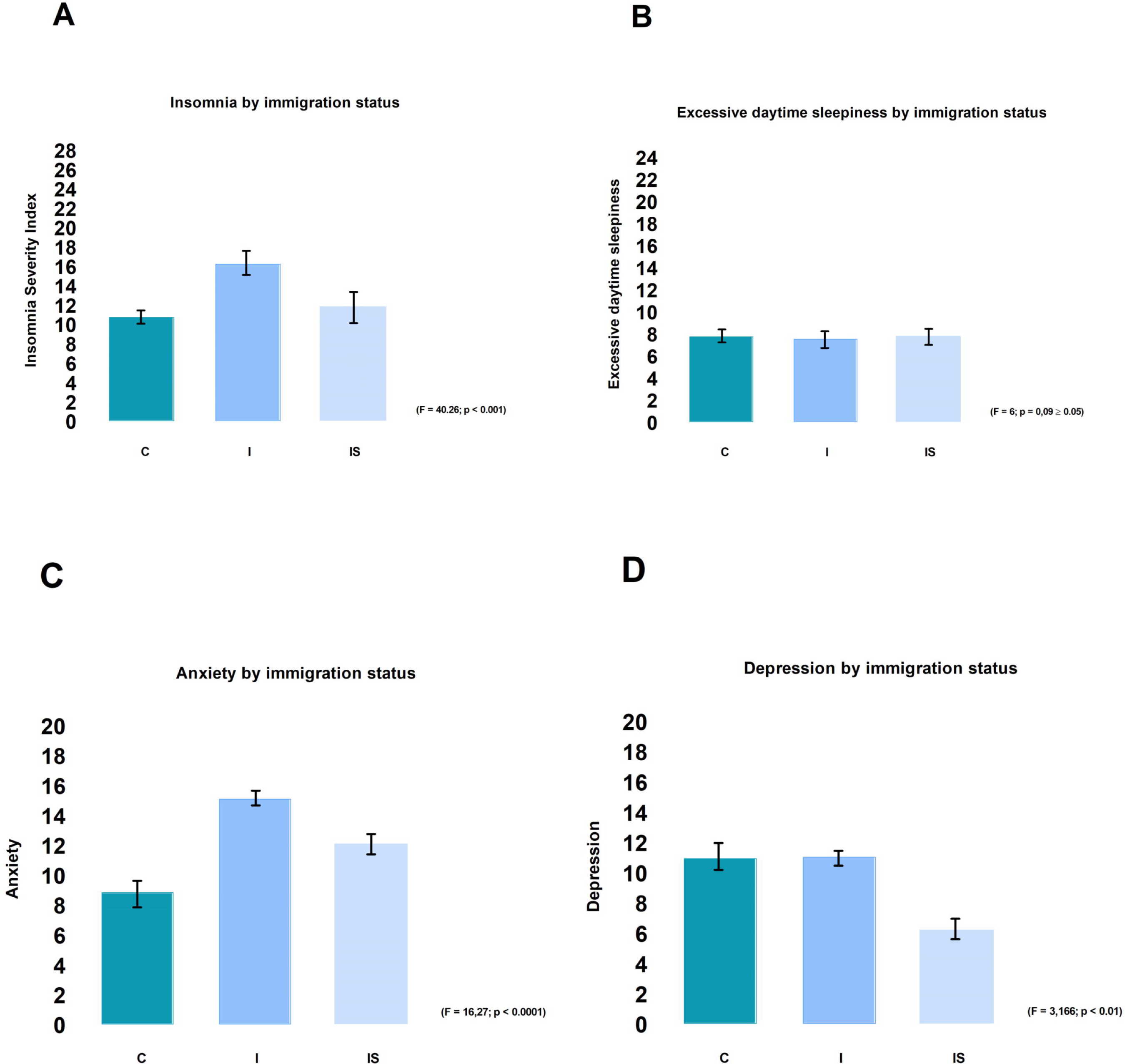
Differences between citizens, international students and immigrants in their neuropsychiatric disorders. A one-way ANOVA with a Tukey multiple comparisons of means was performed to analyze differences between citizens (C), international students (IS) and immigrants (I) in their level of anxiety, depression, insomnia and excessive daytime sleepiness. Confidence interval:95%, α:0.05

## 4. DISCUSSION

The present research was led to document global health of employees working in the customer service field. More specifically, the present paper is the first round of a pilot study which has objectives to (1) document presence of sleep disorders among customer service advisors (2) document presence of anxiety and depression in this population (3) determine the influence of the socioeconomic status, duration in position and full time or part-time shift on the diseases above. The results revealed that people working in customer service are aged 26 ± 4 years old for men and 21 ± 2 years old for women, with an undergraduate or a secondary/high school degree. The majority worked full time, are single and have reported at least two of the neuropsychiatric disorders assessed in the present study. Among customer service employees enrolled in this study, all neuropsychiatric disorders investigated were present and significantly higher for those working full time. Perceived socioeconomic status (pSES) was almost similar for full time and part time workers with a mean score of 4.8 on the MacArthur scale of subjective social status. Results also revealed that duration in position was an excellent predictor of insomnia, excessive daytime sleepiness and anxiety (respectively with R^2^=91,83%, R^2^=81,23% and R^2^=87,46%) but a moderate predictor of depression (R^2^=69,14%). The pSES was a moderate predictor of sleep disorders (respectively R^2^=62,04% for insomnia and R^2^=53,62% for excessive daytime sleepiness) but had a strong association with anxiety and depression (R^2^=82,95% for anxiety and R^2^=89,77% for depression). Then, it was found that insomnia and anxiety are more prevalent for immigrants and international students compared to Canadians, while depression was similarly higher for Canadian and immigrants compared to international students. Finally, it was found that excessive daytime sleepiness has the same trend in the three subgroups. Considering these results above, the three objectives were completed. First, insomnia and excessive daytime sleepiness were found in the sample, with statistic for men and women on one side; and for part time and full-time workers on the other side. No previous study reporting these sleep disorders for this specific population was found until now in the literature. It is one of the main strengths of this paper. Data were reported about cardiovascular and metabolic diseases of people working in sector with rotating shift (Kalmbach et al., 2015) and some others showed side effects of irregular shift on allostatic load and mood disorders (Touitou et al., 1990; Kalmbach et al., 2015; Travis et al., 2016). These studies reported that some work with rotating shifts like oil refinery operators or nurses, were associated to changes in the metabolism hormones like melatonin, prolactin, testosterone and cortisol without any apparent phase shift of these hormones (Touitou et al., 1990). Further evidence also demonstrated that diurnal rhythms in cortisol, melatonin and HRV are not adapted to night work after 1-3 consecutive night shifts (Jensen et al., 2016). When a worker experimented during several months an irregular work shift, he finally get a circadian disruption which was recently recognized a risk factor for diabete, cancer and cardiovascular outcomes (Straif et al., 2007; Jensen et al., 2016; Travis et al., 2016). Additional evidence has also linked shift work with sleep disorders, with a proportion estimated by the 2005 International Classification of Sleep Disorders around 2 to 5% of night shift workers (Boivin and Boudreau, 2014). The findings of this paper confirmed the impact of shift work on sleep, by adding customer service work in the list of work associated to sleep disorders. It is in line with previous studies which stated that, individual tolerance to irregular shift work was a complex issue influenced by the duration in position, the chronotype preference and the genetic background (Boivin and Boudreau, 2014; Jensen et al., 2016; Jagannath et al., 2017). Few sleep disorders like excessive daytime sleepiness appear during intermittent night shifts and they are related to desynchronization of the circadian rhythms, disturbing attention and vigilance of the workers after an unknown period (Boivin and Boudreau, 2014; Jensen et al., 2016). Obviously, the resulting shift work sleep-wake disorder affects the quality of life of the worker by creating cognitive decline or mood disorders (Kalmbach et al., 2015). Regarding anxiety and depression, few studies reported influence of type of work and type of shift on neuropsychiatric outcomes (Averina et al., 2005; Harris et al., 2010). The present study is the first which reports prevalence and proportion of anxiety and depression in a population of customer service employees, and also confirms the existence of a strong relation between socioeconomic status and mood disorders in line with few articles reporting possible relation between socioeconomic status and health (Mulatu and Schooler, 2002; Averina et al., 2005; Phelan et al., 2010; Hawkley et al., 2011; McEwen, 2017; Etindele Sosso et al., 2018a; Etindele Sosso et al., 2018b; Etindele Sosso and Papadopoulos, 2018; Etindele et al., 2019; Sosso et al., 2019). The present study alerts on the potential effect of working full time in a call center as a risk factor for neuropsychiatric illnesses. The findings of this study clearly demonstrated that people working full time obtained the higher score in the different questionnaire assessing anxiety, depression, insomnia and excessive daytime sleepiness. Higher score means bad diagnosis for all the diseases evaluated, and even if the current transversal design did not allow to follow the impact of working full time longitudinally on sleep disorders, anxiety and depression; findings presented here are trustworthy pictures of the mental health of the participants who are mainly customer service advisors. Taking in account presence of rotating shift and irregular shift work in customer service, these findings document an existing association between work in customer service and neuropsychiatric diseases. The results also question the role of the duration in position and socioeconomic status as risk factors for neuropsychiatric diseases for customer service employees. While social factors and sociodemographic factors may influence health in general (Adler et al., 2000; Contoyannis and Jones, 2004; Chen et al., 2005; Assari et al., 2016; Etindele Sosso and Papadopoulos, 2018; Sosso et al., 2019) and even if evidence exists on their key role as mediators between socioeconomic status and health (MULATU and SCHOOLER, 1999; Contoyannis and Jones, 2004; Phelan et al., 2010; Hawkley et al., 2011; Chen et al., 2013; El-Sheikh et al., 2013; Grandner et al., 2013); nobody really investigated how people working in such field are exposed to chronic stress, and current literature contains no data on employee’s health of this sector. The present paper draws an interesting profile in terms of age, education, socioeconomic status, immigration status and duration in position; of people working in customer service in Canada. This is another strength of the present article, because no other study examined so deeply this population who is probably also exposed to shift work sleep-wake disorder and many others neurological disorders commonly found in similar works (Boivin and Boudreau, 2014; Jensen et al., 2016). Despite previous questions raised by these results and debated in the previous paragraphs, some limitations exist for this paper. First, the difficulty to know what the exact role of each participant in their respective companies are. This may be a limit because the level of stress experienced by a manager is different from the one experienced by an advisor, as showed by few authors who documented the difference in daily stress of cortisol secretion due to some socioeconomic markers like education, occupation or income (Harris et al., 2010; Hawkley et al., 2011; Green et al., 2014). In other words, someone with a high socioeconomic status like a manager or a medical doctor will have most often less stress and low level of cortisol compared with an individual with a low socioeconomic status like a garbage collector or a delivery man (MULATU and SCHOOLER, 1999; Karlamangla et al., 2013; Stawski et al., 2013) and the present study did not investigate this difference. More research is necessary to produce more details about the influence of an individual’s position in a company on his health outcome. Another limit found here was the absence of medical history or confounding factors like weight and presence of respiratory disease. It was reported that excessive daytime sleepiness is often associated to sleep apnea which can be mistaken for with snoring, which is strongly associated with obesity (Lack and Sweetman, 2016; Tsai, 2016). Some treatments for anxiety may induce insomnia (Lack and Sweetman, 2016) and some participants taking these kinds of medication may have been included in this study. Even if this has no influence on the study’s objectives and results, next studies should pay attention on this possible bias which may affect research where mental disorders, or any other diseases studied are linked to medical history or can be influenced by medication. In the same order, subscales of the questionnaire were not analyzed in this case only because it was not necessary for this pilot study; but it will be an improvement if further research investigates the current findings to see if the same trends arise when looking at sleep quality, sleep satisfaction or other components based on a different sleep questionnaire such as PSQI.

In conclusion, customer service employees are exposed to a continuous stimulation of their cognitive functions in addition to different stressors which can progressively and silently damage the nervous system. Investigations on mental and physical health of customer service advisors are worthy of interest, and understanding how their work, their rotating shifts and their socioeconomic status influence their resilience and their performance at work; may help comprehension of similar health issues emerging in similar populations with similar occupations. For now, here are some practical suggestions to reduce neuropsychiatric disorders for customer service employees or at least mitigate the work burden on their brain:

### Avoid rotating shift as much as possible

Like explained previously, the stabilization of an individual circadian rhythm is difficult with an irregular sleep and a fast rotating shift (Canuto et al., 2013; Kalmbach et al., 2015). It is better to recruit employees working only night shifts or those working only day shifts. Changing shifts of an individual will create more cost for the employer because of health outcome employees will develop. In the present study, neuropsychiatric diseases were investigated but it may be possible that the same employees have musculoskeletal diseases, metabolic disorders or burn-out; which were already reported as side effects of rotating shift (Canuto et al., 2013; Kalmbach et al., 2015).

### Encourage physical activity during a shift

Many studies reported the benefits of moderate physical activity on the health, whatever type of activity it is (Bancroft et al., 2015; Song et al., 2015; White et al., 2016; Knapp et al., 2018). It is true for elderly, children or adolescents. It is not a waste of money to extend the lunch time during a full shift, to allow people to walk somewhere, do a short yoga session or bike (Bancroft et al., 2015; Song et al., 2015; White et al., 2016; Knapp et al., 2018).

### Increase part-time shift

like demonstrated in this study, people working full time reported much more neuropsychiatric diseases than part time workers. According to the diversity of the canadian population and the different subgroups of participants in this study, the mental disorders found here are not related to ethnicity, age or some bias; the sample is very representative of the general population. This also means that results are clearly a warning on the dangers related to the to the stressful environment that is customer service. In my opinion, reducing full shifts to 6 or 7 hours including a 30 minute-lunch break may be a good compromise for both the employers and the employees in terms of cost, to decrease the number of sick day-related absences and to increase work performance

### Enlarge the investigation to other countries

Canadian population is similar because of immigration to other countries like United Kingdom, United State of America, France, Belgium, Australia and Germany. Customer service is an important part of their economy as industrialized countries, so, without knowing it, these countries probably face the same neuropsychiatric disorder problems with their employees. More research in these countries may be interesting and informative.

## Data Availability

All data are available on demand.

## Acknowledgments

This work was possible with the participation and collaboration of Mr Shaun Thivierge, Mr Antoine Bonnet and Ms. Caroline Courtemanche; respectively team leader at ICT Marketing, manager at Public Mobile Telecom and recruiter for iOS Center. Many thank to Ms. Audrey-Anne Charbonneau who explained to us federal rules and laws related to ccustomer service in Canada, and who shares electronic link of the study with the managers /administrators of 6 call centers. Thanks to Dr Paul Rodrigues and Jill Vandermeerschen who revised statistical analysis and writing of the section materials & methods. Thanks to Dr Giuseppe Curcio and Dr Dimitrios Papadopoulos for their critical and useful comments on the final form of this manuscript. Thanks to Tommy Khoury who performed language proofreading during the writing of this paper.

## Author Contributions Statement

FA, ES collected the data, analyzed data and wrote the manuscript in his present form.

